# Passive Microwave Radiometry (MWR) for diagnostics of COVID-19 lung complications in Kyrgyzstan

**DOI:** 10.1101/2020.09.29.20202598

**Authors:** Batyr Osmonov, Lev Ovchinnikov, Christopher Galazis, Berik Emilov, Mustafa Karaibragimov, Meder Seitov, Sergey Vesnin, Chingiz Mustafin, Turat Kasymbekov, Igor Goryanin

**Affiliations:** Educational - scientific medical center of Kyrgyz State Medical Academy (KSMA) named after I.K. Ahunbaev, Kyrgyzstan; Medical Microwave Radiometry LTD, UK; University of Edinburgh, UK; International Medical University (IMU), Kyrgyzstan; Komfort Clinic, Kyrgyzstan

**Keywords:** COVID-19, Passive Microwave Radiometry (MWR), Infrared Radiometry (IR), RT-PCR, CT

## Abstract

It becomes clear that the COVID-19 virus is spreading globally due to limited access to diagnostics tests and equipment. Now, most of the diagnostics has been focused on RT-PCR, chest CT manifestations of COVID-19. However, there are problems with CT due to infection control, lack of CT availability in LMIC (Low Middle Income Countries) and sensitivity of RT-PCR. Passive microwave radiometry (MWR) is a cheap, non-radioactive and portable technology. It has already been used for diagnostics of cancer, and other diseases. We have tested if MWR could be used for early diagnostics of pulmonary COVID-19 complications. This was a randomized controlled trial to evaluate MWR in patients with COVID-19 pneumonia in hospitals, and in healthy individuals. We have measured skin and internal temperature at 30 points on both lungs. Pneumonia and lung damage were diagnosed by CT scan and doctor diagnosis (pn+/pn-). COVID-19 was determined by RT-PCR tests (covid+/covid-). The best MWR results were obtained between pn-/covid- and pn+/covid+ groups with sensitivity 92% and specificity 75%. The study suggests that the MWR is a safe method for diagnostics of pneumonia in COVID-19 patients. Since MWR is an inexpensive, it will ease the financial burden for both patients and the countries.

**Clinical Trial Number:** NCT04568525

**Study design:** Randomized controlled clinical trials

## 1. Introduction

A significant number of deaths occurred in COVID-19 patients with multiple concomitant diseases, such as interstitial pneumonia, acute respiratory distress syndrome, and subsequent multiple organ failure [1]. Although severe lung damage has been described at any age, in some people at high risk, the virus is more likely to cause complications. In affected persons various degrees of dyspnea and radiological signs are observed [2, 3]. At this moment most of the research was focused on CT manifestations of the chest COVID-19 [4, 5]. Ground-glass opacities in the early stage, paving patterns and diffuse damage in the later stage. In contrast to the great sensitivity of chest CT, the specificity was relatively low with reporting about 25–33%. CT patterns are observed in other pneumonia and non-infectious inflammatory lung diseases but in a pandemic context might harbor diagnostic potential for COVID-19 infection especially for patient triage [9]. In addition, there are obstacles of using CT due to the infectious controls related to patient transportation, as well as disinfection of CT rooms after examining the patient and the lack of accessibility. Portable chest x-ray could be used to minimize risk of infection [6]. In contrast to the great sensitivity of chest CT, the specificity was relatively low with reporting about 25–33% In the early stage ground-glass opacities are the predominant lesion. In the next stage, crazy paving patterns mark the inflammation. Peak stage is marked by fibrosis and diffuse damage.

These CT lesions are also observed in other pneumonia and non-infectious inflammatory lung diseases but in a pandemic context might harbor diagnostic potential for COVID-19 infection especially for patient triage.[9] In health care settings with limited PCR capacity and long turnaround times, chest CT was proposed as alternative for COVID-19 diagnosis. Studies supporting chest CT as first-line diagnostic tool for COVID-19 showed several methodological concerns [10-12]. There are associated cost and procedural risks of CT [13-15] These lesions are also observed in other pneumonia and non-infectious inflammatory lung diseases. In health care especially with limited PCR and CT.

The other method for COVID-19 diagnosis, RT-PCR, has a variable sensitivity as low as 70% [16]. Specificity of viral swabs in clinical practice varies depending on the site and quality of sampling. In one study, sensitivity of RT-PCR in 205 patients varied, at 93% for broncho-alveolar lavage, 72% for sputum, 63% for nasal swabs, and only 32% for throat swabs. The test results are also likely to vary depending on stage and degree of viral load or clearance [17-19]. Specificity of between 2% and 29% (equating to sensitivity of 71-98%), based on negative PCR tests which were positive on repeat testing. The use of repeat RT-PCR testing as standard is likely to address probable low specificity, and the true rate of false negatives, because not all patients received repeat testing.

Non-expensive, another safer method is required to replace and/or compliment CT and PCR tests. These circumstances make us look for new diagnostic methods.

Passive microwave radiometry (MWR) is a cheap, non-radioactive and portable technology [20]. It has already been used for early diagnosis of cancer, and other diseases. It implies measuring temperature of tissue region by measuring black body emission in microwave range. This allows it to receive signals from subcutaneous tissues hence revealing microwave (internal) temperature changes up to 5 cm deep under the skin. The increase of microwave emission is caused by inflammation, while decrease is caused by fibrosis. The theoretical advantage of MWR is that the temperature manifestations can be revealed before any structural changes can be registered.

The RTM-01-RES device (Fig.1) is a unique commercially available CE marked device. It is registered in UK MHRA MDN 40802 as a microwave thermography system for clinical studies. The device is already registered in Kyrgyzstan for breast cancer diagnostics. During the 1980-90s there were several works on identification of excess microwave emission due to fluid in lungs (on phantoms) which could be indication of inflammatory processes, pneumonia, cancer and other lung disorders. [21,22]. Later results were confirmed by clinical studies for lung cancer [23,24].

**Figure 1.**
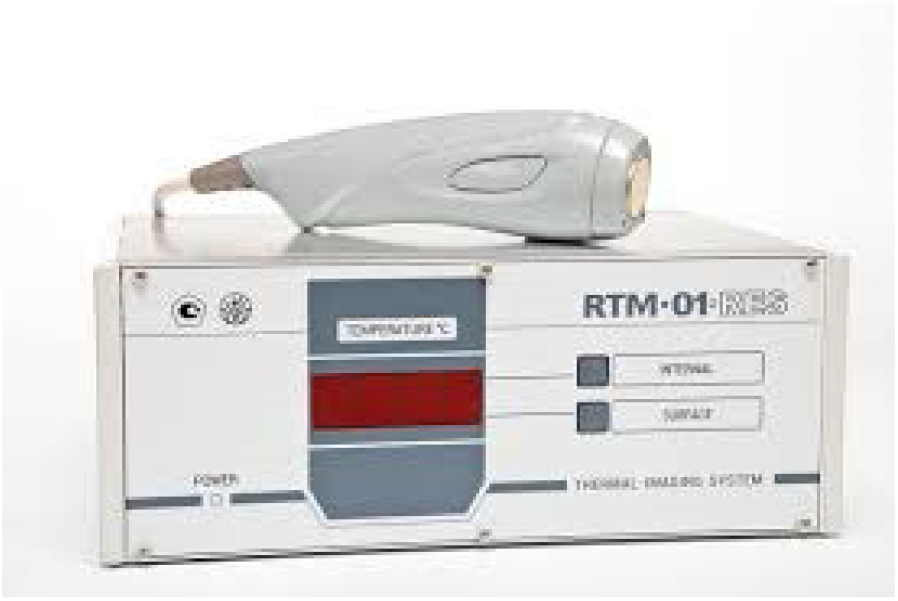
MWR Device. RTM-01-RES.

The purpose of this study was to investigate the value of MWR and compare microwave emission from left and right lungs with chest CT and RT-PCR to determine its diagnostic performance in individuals with COVID-19 symptoms.

## 2. Materials and Methods

In this trial (Kyrgyz Committee Clinical Trial Number: 01-2/141 27 May 2020), from June, 1 2020 to August, 1 2020, we performed parallel MWR, PCR and CT tests, for individuals with COVID-19 admitted to the hospital for medical emergencies related to COVID-19 and pneumonia suspicion. Siemens Ecoline CT scanner, and HITACHI, Radnext 50 Chest X-Ray was used. RT-PCR test were done using “DNA technology” kits https://www.dna-technology.ru). For MWR and IR measurements RTM-01-RES was used MMWR LTD, UK (www.mmwr.co.uk)

This is an analysis of a single-center prospective trial on consecutive individuals admitted to Medical center of KSMA and BICARD clinic from July 1, 2020 to August 1, 2020. KSMA is a central-network regional hospital that provides tertiary health care for a community of 500,000 inhabitants. All individuals admitted to the hospital with clinical suspicion of COVID-19 pneumonia (confirmed by experienced pulmonologists, so ‘symptomatic individuals,’ inclusion criteria), received a combined screening with chest CT and RT-PCR. We used the COVID-19 case definition as specified by the World Health Organization (WHO) document [25] for classifying symptomatic individuals. Lung comorbidity, individuals without COVID-19 symptoms did not receive chest CT. Exclusion criteria include lung comorbidity, like exacerbation of COPD, very severe COPD with hypoxia (FEV1 <40%, saturation <92% at an altitude of 760 m), co-morbidities, such as cardiovascular diseases, i.e. unstable systemic arterial hypertension, coronary heart disease; stroke; sleep apnea; pneumothorax last 2 months, neurological, rheumatological or psychiatric illnesses, including excessive smoking (> 20 cigarettes per day), kidney failure.

The study was approved by the Kyrgyz Republic Review Board, and informed consent was obtained from all subjects. Overall, we have measured internal (MWR) and skin (IR) temperature on 195 subjects, 74 male and 121 females from 18 to 75 years old (Table 1). 149 of them were hospitalized with pneumonia symptoms to Medical center of KSMA and BICARD clinic Bishkek, and 46 was chosen from healthy volunteers with no COVID-19 and pneumonia symptoms. RT-PCR test was performed for each hospitalized subject, 116 subjects found positive.

**Table 1.**
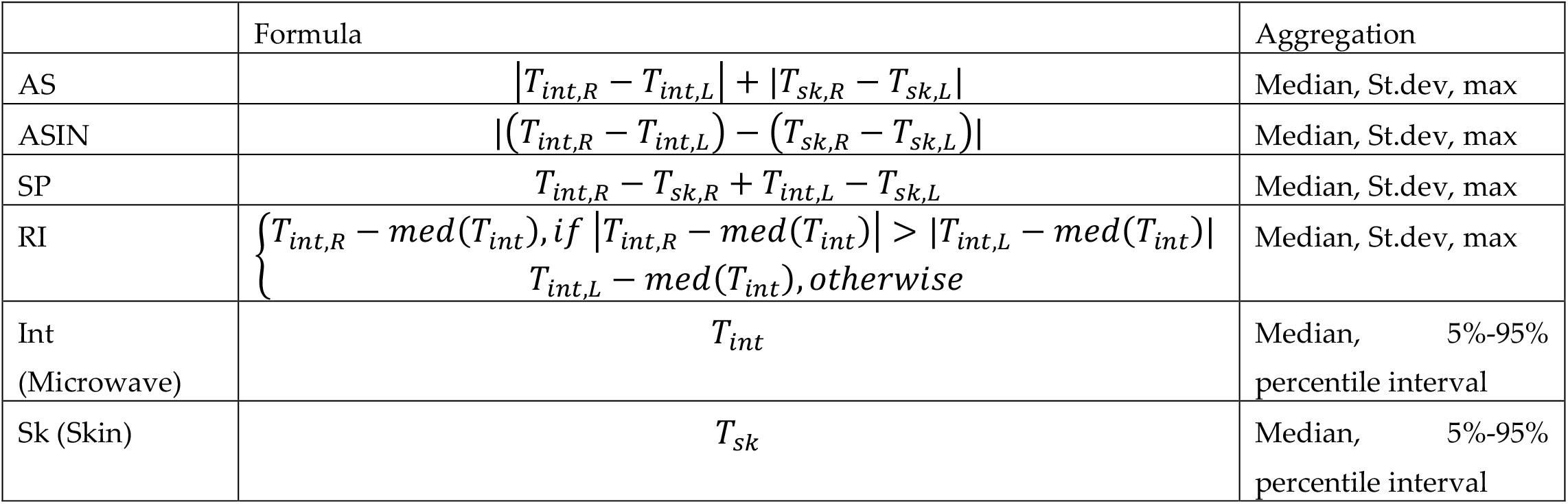
Aggregation metrics.

Healthy volunteers without COVID-19 and pneumonia symptoms did not receive chest CT. but they were exposed to MWR scan. CT scans were performed with Siemens Ecoline CT scanner and described in a standard routine by hospital physician, who wasn’t directly involved in this research. RT-PCR test were done using “DNA technology” kits https://www.dna-technology.ru/). For MWR and IR measurements RTM-01-RES (Figure 1) was used (MMWR LTD, UK www.mmwr.co.uk). BMI, auxiliary (armpit) temperature and SpO2 were additionally assessed as a part of hospital submission routines. 93 of 195 subjects were measured through thin clothes, 102 measured without clothes.

Novel MWR measurement technique was designed for this research. There were 30 points on a body, 28 symmetrical (R1-L14) and two additional (T1 and T2), as shown on Fig. 2-4. Some of patients were measured with thin clothes on, and some were measured without clothes in a stand position.

**Figure 2:**
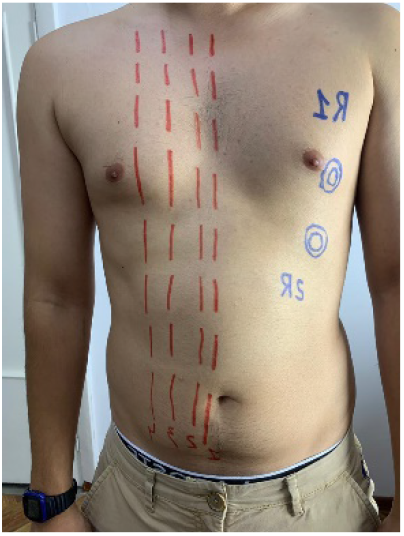
Right Lung R1 R2 measurements points.

Software application “RTM Diagnosis 1.79” was configured to perform these measurements, which can gather data from device and plot microwave mages.

### Statistical Approach

The data was analyzed patient-wise. As variables, 30 internal (MWR) temperatures and 30 skin (IR) temperatures were collected. As a reference, two conditions were separately used – confirmed diagnosis of pneumonia on CT (pn+, pn-) and positive RT-PCR test (covid+, covid-). So, there were 4 groups. (covid+/pn+), (covid-/pn-), (covid+/pn-), and (covid-/pn+), and statistical analysis was aimed to tell if MWR can predict subject being in specific group.

To estimate possible sensitivity and specificity we build simple classifier based on k-neighbors strategy. Phase space is built from metrics which shows to have significant differences between groups. Then, we split the data in 3:1 to ratio and predict if patient in specific group, based on remaining data. This procedure is repeated for 1000 times to eliminate randomness from splitting the data. Distribution of sensitivity and specificity is plotted as a random value.

## 3. Results

### 3.1 Clinical results

RT-PCR test was performed for each hospitalized subject, 116 subjects found positive. CT scans of 89 patients were diagnosed with pneumonia. 87 of 89 patients had bilateral pneumonia. Pneumonia (CT) and RT-PCR results were set as reference variables. Four groups were split based on these two variables. Ambient temperature varied from 27 to 30. Average BMI for all subjects is 26.3 (Table 2) SpO2 was measured for each subject while initial examination at hospital. Additionally, SpO2 and auxiliary (armpit) temperature was measured for each subject while initial examination at hospital Within 24h from admission all individuals were imaged by CT. Radiologists with many years of experience reviewed the CT test, and assessed left and right % of lungs damage. They were blinded to (a)symptomatic status and RT-PCR result. Median lung damage percentage for subjects with positive RT-PCR test is 40%. Auxiliary (armpit) temperature was measured for each subject while initial examination. Median axillary temperature is 36.6 for covid-group, and 36.7 for covid+ group. MWR temperature measurements were done in lung projection, as shown in Figure 2, 3, 4, in symmetrical points pairwise, in 30 positions total. All data are in Supplementary 1 xlsx file.

**Table 2.**
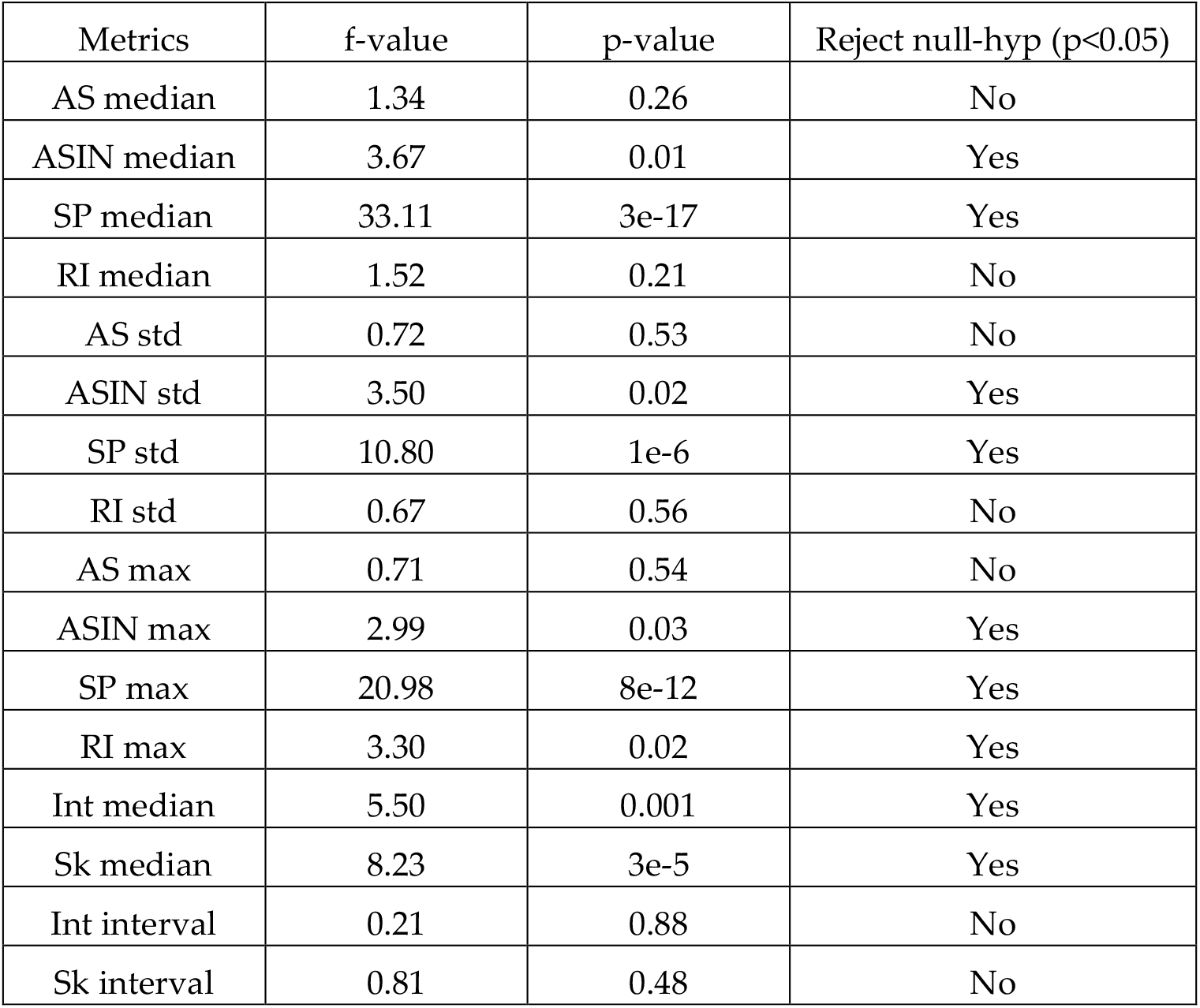
ANOVA statistics.

| Metrics | f-value | p-value | Reject null-hyp ($p < 0.05$) |
| --- | --- | --- | --- |
| AS median | 1.34 | 0.26 | No |
| ASIN median | 3.67 | 0.01 | Yes |
| SP median | 33.11 | 3e-17 | Yes |
| RI median | 1.52 | 0.21 | No |
| AS std | 0.72 | 0.53 | No |
| ASIN std | 3.50 | 0.02 | Yes |
| SP std | 10.80 | 1e-6 | Yes |
| RI std | 0.67 | 0.56 | No |
| AS max | 0.71 | 0.54 | No |
| ASIN max | 2.99 | 0.03 | Yes |
| SP max | 20.98 | 8e-12 | Yes |
| RI max | 3.30 | 0.02 | Yes |
| Int median | 5.50 | 0.001 | Yes |
| Sk median | 8.23 | 3e-5 | Yes |
| Int interval | 0.21 | 0.88 | No |
| Sk interval | 0.81 | 0.48 | No |

**Figure 3:**
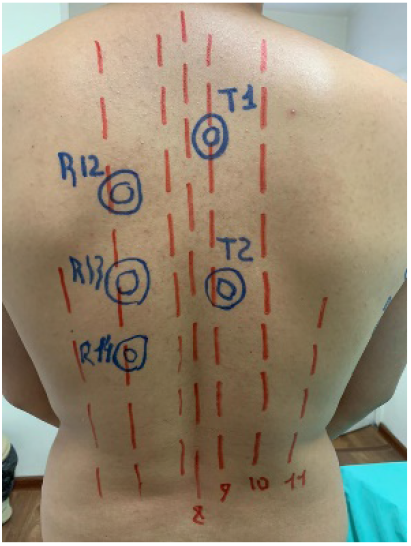
Right Lung T1 T2 R12 R13 R14 measurements points.

**Figure 4:**
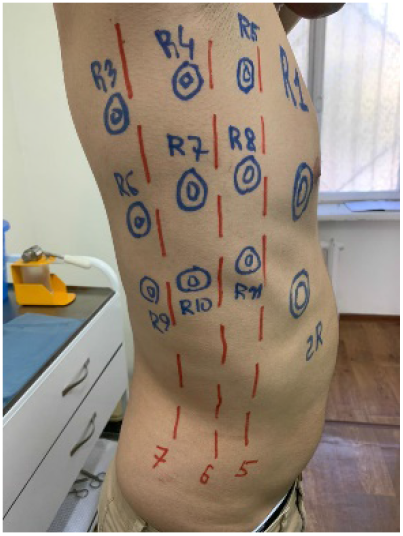
Right Lung R3-R11 measurements points.

### 3.2 Clinical images

Fig. 5 shows typical Microwave (MWR) Image of COVID-19 pneumonia. On both left and right lung, one could see a large internal temperature difference in blue areas (low temperature due to fibrosis and red areas (high temperature due to inflammation.

**Figure 5.**
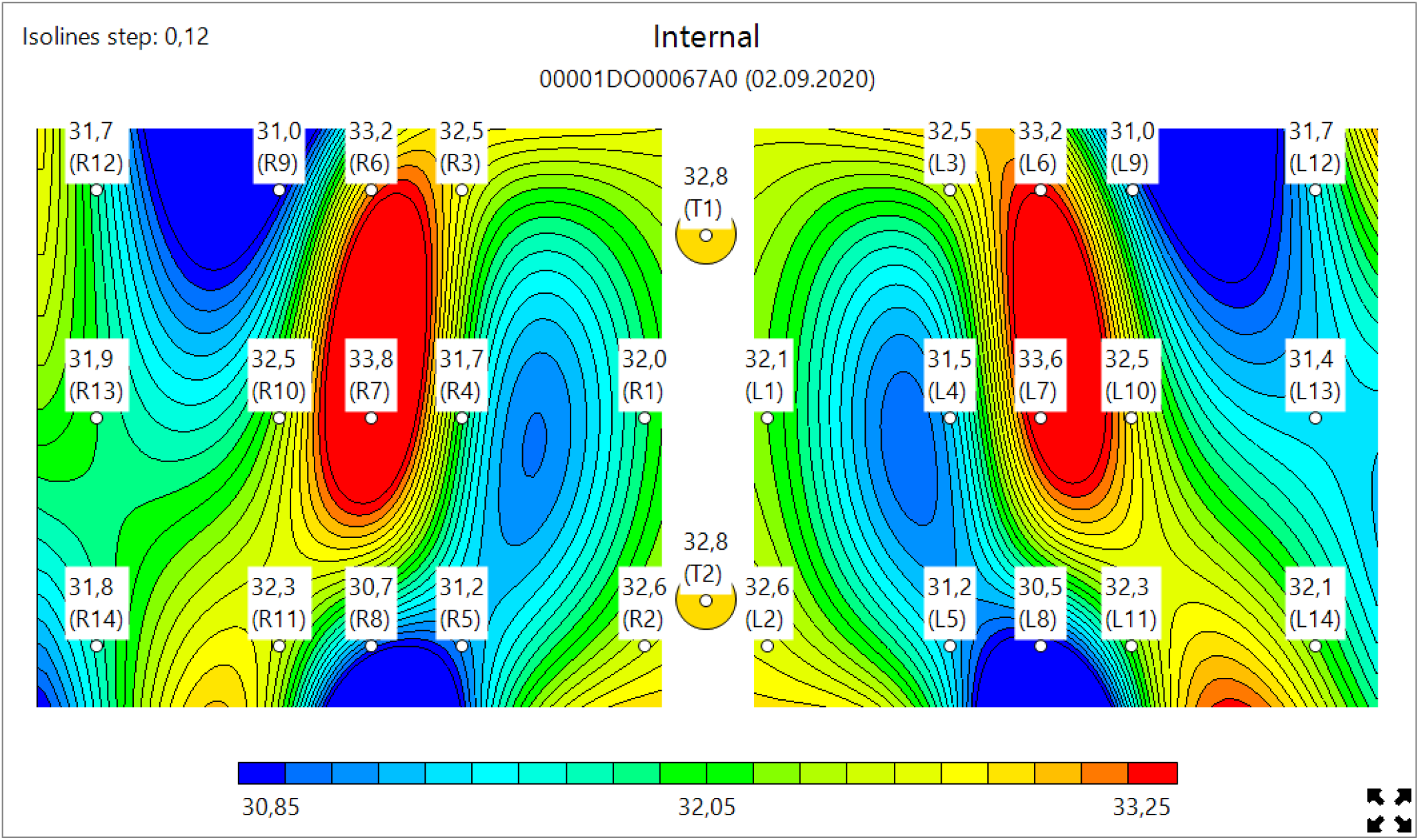
Typical Microwave Image. COVID-19. Left and Right Lungs. Large internal temperature difference. Blue (low temperature due to Fibrosys) and Red (high temperature due to inflammation

Fig. 6 shows lungs of healthy individual, and on Fig 7 depicted nob COVID-19 pneumonia where regions with inflammations only could be observed, boy no blue zoned are visible.

**Figure 6.**
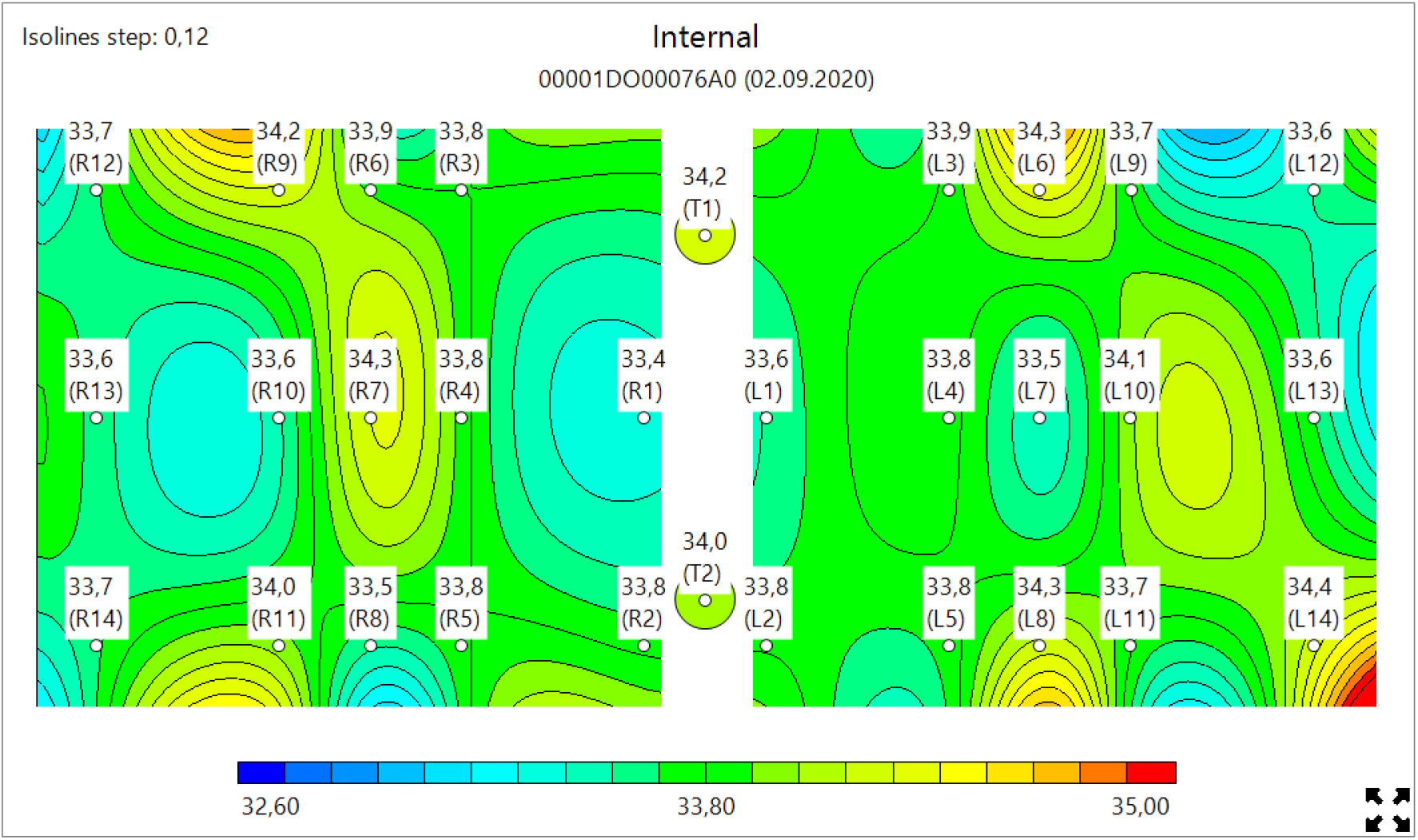
Typical Microwave Image. Healthy Lungs. No Blue or Red areas.

**Figure 7.**
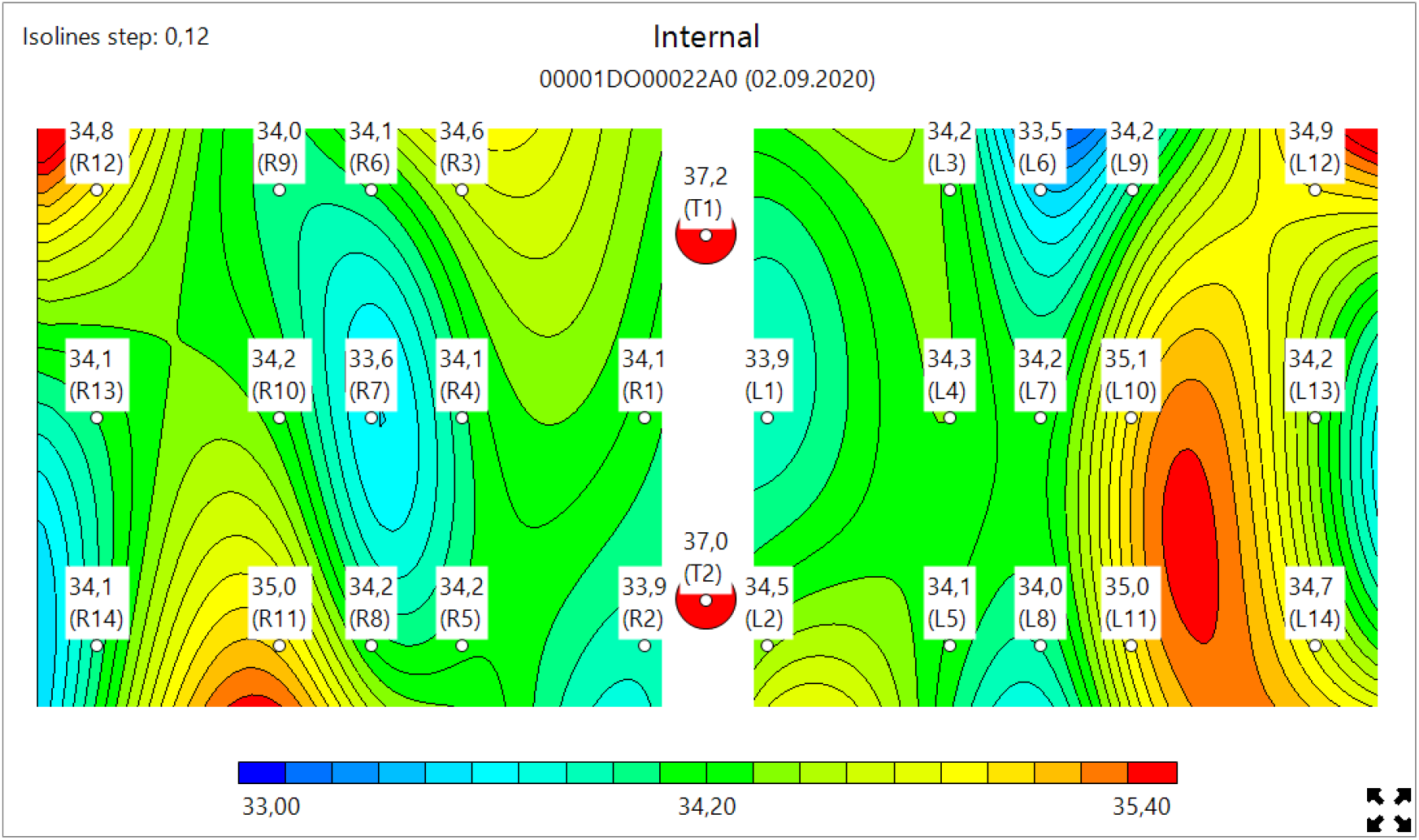
Typical Microwave Image. NON COVID-19 pneumonia. Red (Inflammation area only in the left Lung. No Blue areas due to fibrosis in both lungs.

### 3.3 Statistical results

It was found that internal (MWR) and skin (IR) temperature averages separately will not give a full diagnostic power. So, the following integral metrics were calculated for each subject (Table 1), where T_(int,R), and T_(int,L) is the internal (MWR) temperature of right and left lungs, T_(sk,R) T_(sk,L) is the skin (IR) temperature of right and left lungs, med is a median for corresponding dataset (all 30 points). Each metric was separately tested to have statistically significant differences between 4 groups. (covid+/pn+), (covid-/pn-), (covid+/pn-), and (covid-/pn+).

First, MWR variables for each patient were transformed into several integral metrics which is described in Table 1.

Then, each of those metrics is separately subjected to one-way multi-group ANOVA (Table 2) to check if 4 groups are likely to have identical mean. For metrics having p < 0.05, pairwise Tukey test (Table 3) was performed to assess difference in metrics between groups pairwise. Those assessments were done using python script (scipy.stats.f_oneway, statsmodels.stats.multicomp.pairwise_tukeyhsd [26, 27]. Appendix B

**Table 3.** Significant pairs.

| Criteria | Significant pairs |
| --- | --- |
| ASIN median | Covid- pneumonia- and Covid- pneumonia+ (diff = +0.12, $p = 0.04$)<br>Covid- pneumonia- and Covid+ pneumonia+ (diff = +0.12, $p = 0.02$) |
| ASIN std | Covid- pneumonia- and Covid+ pneumonia+ (diff = +0.4, $p < 0.001$) |
| SP median | Covid- pneumonia- and Covid- pneumonia+ (diff = +1.7, $p < 0.001$)<br>Covid- pneumonia- and Covid+ pneumonia- (diff = +1.2, $p < 0.001$)<br>Covid- pneumonia- and Covid+ pneumonia+ (diff = +2.4, $p < 0.001$)<br>Covid+ pneumonia- and Covid+ pneumonia+ (diff = +1.2, $p < 0.001$) |
| SP std | Covid- pneumonia- and Covid- pneumonia+ (diff = +0.9, $p < 0.001$)<br>Covid- pneumonia- and Covid+ pneumonia+ (diff = +0.8, $p < 0.001$)<br>Covid- pneumonia+ and Covid+ pneumonia- (diff = -0.6, $p = 0.01$)<br>Covid+ pneumonia- and Covid+ pneumonia+ (diff = +0.5, $p = 0.01$) |
| SP max | • Covid- pneumonia- and Covid- pneumonia+ (diff = +4.42, p < 0.001)<br>• Covid- pneumonia- and Covid+ pneumonia- (diff = +1.95, p = 0.005)<br>• Covid- pneumonia- and Covid+ pneumonia+ (diff = +3.94, p < 0.001)<br>• Covid- pneumonia+ and Covid+ pneumonia- (diff = -2.46, p < 0.001)<br>• Covid+ pneumonia- and Covid+ pneumonia+ (diff = 2.0, p = 0.002) |
| RI max | • Covid- pneumonia- and Covid- pneumonia+ (diff = +0.6, p = 0.03) |
| Int median | • Covid- pneumonia- and Covid- pneumonia+ (diff = +1.3, p = 0.011)<br>• Covid- pneumonia+ and Covid+ pneumonia- (diff = -0.7, p = 0.002)<br>• Covid+ pneumonia- and Covid+ pneumonia+ (diff = +0.5, p = 0.03) |
| Sk median | • Covid- pneumonia- and Covid+ pneumonia- (diff = -0.8, p < 0.001)<br>• Covid- pneumonia- and Covid+ pneumonia+ (diff = -0.9, p < 0.001)<br>• Covid- pneumonia+ and Covid+ pneumonia+ (diff = -0.6, p = 0.04) |

From ANOVA it was found that the following criteria are to have differences between groups: ASIN (all aggregates), SP (all aggregates), RI max, Int median, Sk median.

From pairwise Tukey test (Table 3) it was found that border groups (covid-/pn-and covid+/pn+) differ for almost all key metrics. COVID-19 patients are characterized by decrease in skin temperature (median from −0.6 to −0.9), and consequently, increase in spread between internal and skin (median from +1.2 to +1.7, peeking up to +2.0 in single points). Pneumonia characterized by relative increase in specific points, median +0.6.

We see a significant difference between 4 groups, and the method has high potential for revealing acute COVID-19 pneumonia state. However, there is marginal efficiency to separate COVID-19 positive patients without symptoms. Method seems to work well on isolated pneumonia, it is more efficient for COVID-19 patients. Overall, MWR could identify pn-/covid- and pn+/covid+ groups with sensitivity 92% and specificity 75%, covid-/covid+ 70% sensitivity 40% specificity, pn-/pn+ 75% sensitivity 72% specificity. (Fig. 8, 9, 10).

**Figure 8.**
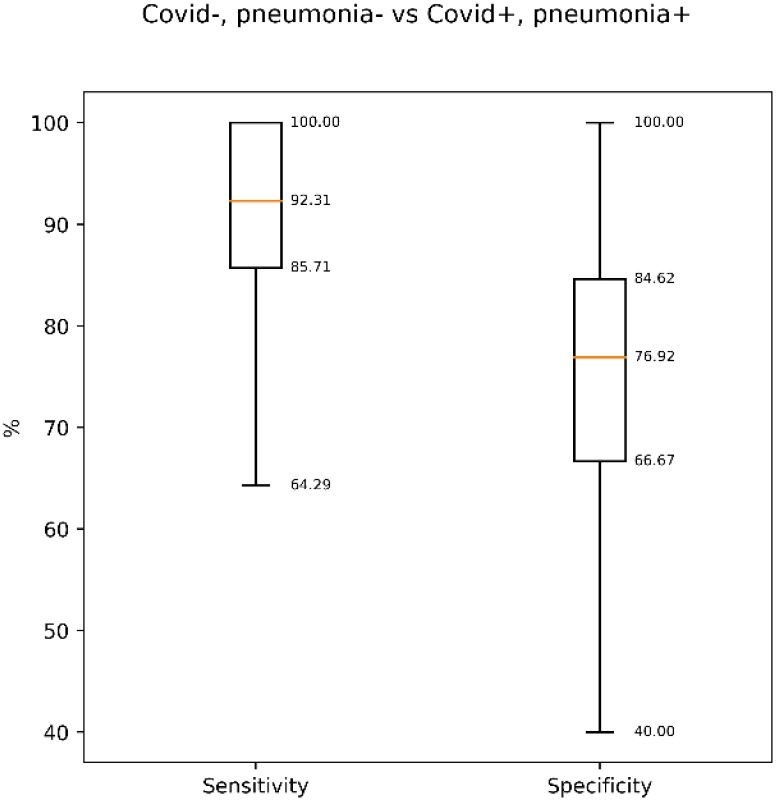
MWR SENSITIVITY AND SPECIFICITY. Covid+/pn+ vs Covid-/pn-groups.

**Figure 9.**
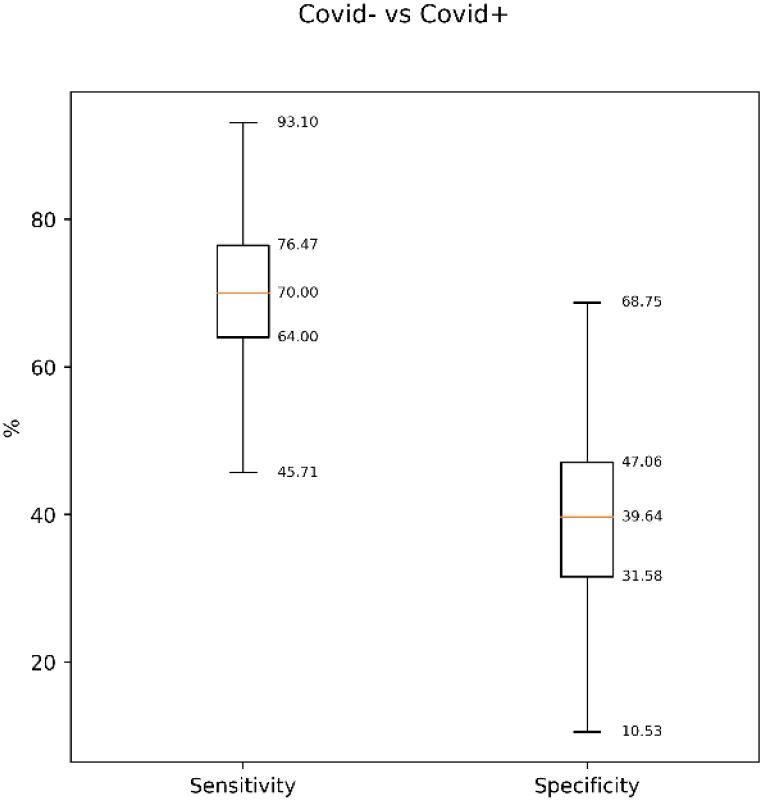
MWR SENSITIVITY AND SPECIFICITY. covid-vs covid+.

**Figure 10.**
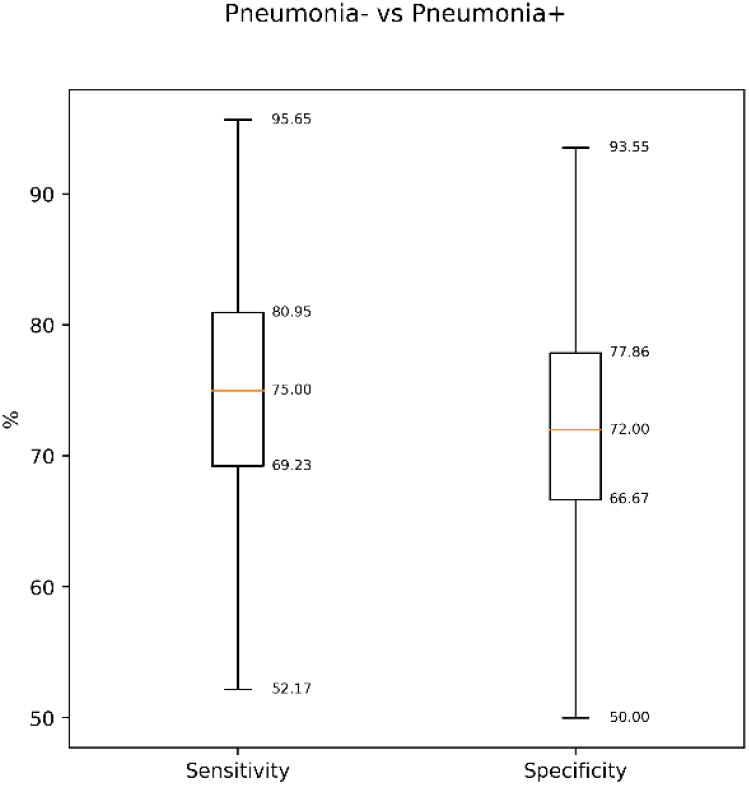
MWR SENSITIVITY AND SPECIFICITY. pn+ vs pn-.

In addition, MWR method has potential for revealing pneumonia states both in COVID- and COVID+ patients. There is observation that skin temperature is lower for COVID+ patients makes hope to use this method to separate COVID-19 pneumonia, however it is not clear yet if this effect is reproducible. It is important to combine internal and skin temperatures to estimate the difference. Otherwise, variations in room temperature and BMI amount will make method ineffective. There were preliminary activities to Deep Neural Network learning system that converts temperature readings and integral metrics to two groups (pn+ and covid+ separately). For pneumonia, it achieves sensitivity 73.91% and specificity 76.47%. For COVID-19, it achieves 68.18% and 72.22% respectively. Details of the process are beyond this paper and will be described and published separately. See [28, 29] for more details.

## 4. Discussion

We aimed to investigate the performance of MWR to diagnose lung complication in SARS-CoV-2 PCR-positive in individuals. Most studies used CT results as positive or negative, often without a clear definition of a positive CT. One large study reported a 97% sensitivity of chest CT for COVID-19 diagnosis but with a poor specificity of 25% [30]. The actual clinical values of a positive CT result to confirm or negative test result strongly depend on disease progression [31]. In recent trials, sensitivity of chest CT was insufficient to exclude SARS-CoV-2 infection which supports the consensus statements that chest CT should not be used as diagnostic test alone [32].

Our data show that our MWR aggregate metrics had good diagnostic performance for COVID-19 pneumonia but cannot replace RT-PCR as diagnostic test. MWR can be used as alternative triage tool in individuals with COVID-19 symptoms and for the screening of asymptomatic SARS-CoV-2 infections The MWR is no radiation, passive technology. It is portable, cheap, and easy to use. MWR can measure internal or Core Body Temperature (CBT), while IR scanners cannot measure CBT of internal organs, but only skin temperature.

Our study has some limitations. It was conducted in a time frame with high rates of SARS-CoV-2 infections and low prevalence of other viral pneumonia. Higher incidence of seasonal respiratory viral infections will likely decrease specificity of MWR. In, healthy individuals are underrepresented in our data set. It is also very important to consider that this methodology will be readily available for LMIC, and that it is even more convenient to use this method at the primary health care level. Primary healthcare is the first line of treatment for patients around the world, and they are the first to contact patients. MWR is a safe method for both doctors and patients, cheap to organize and, most importantly, mobile and simple. The use of MWR will reduce unnecessary costs for CT and X-ray of the lungs for both patients and the state. Microwave radiometry is radiation-free technology, which is portable, cheap and easy to use. The RTM-01-RES device combines both infrared and microwave sensors which allows to outperform existing IR cameras.

The system could be used for early lung diagnostics more widely where access to CT/PCR is limited, including but not limited to

- Nursery homes
- Ships
- Remote locations (highlands, islands, deserts)
- Board security as complementary to IR
- Detention centers

It was more evidence that COVID-19 could damage the brain, heart, gut and other organs. MWR is already being used for diagnostics of different diseases [20]. It could be used for full body scan, including head (brain), wrist (cardiovascular), lung (respiratory), and guts (GI) to assess organ’s damage and eliminate risks in the COVID-19 rehabilitation stage. In the future, to improve the sensitivity and specificity it would be beneficial to use the same Deep Neural Network (DNN) we have earlier applied for breast cancer diagnostics [28,29], but more data are required.

## 5. Conclusions

This study suggests that the use of MWR is a convenient and safe method for screening diagnostics in COVID-19 patients with suspected pneumonia. Since MWR is inexpensive, it will ease the financial burden for both patients and the countries, especially in LMIC

## Data Availability

The xls file with blinded clinical data is attached in supplementary materials.

## Appendix A

### LUNG DAMAGE ASSESMENT

**Appendix A. Figure 1.**
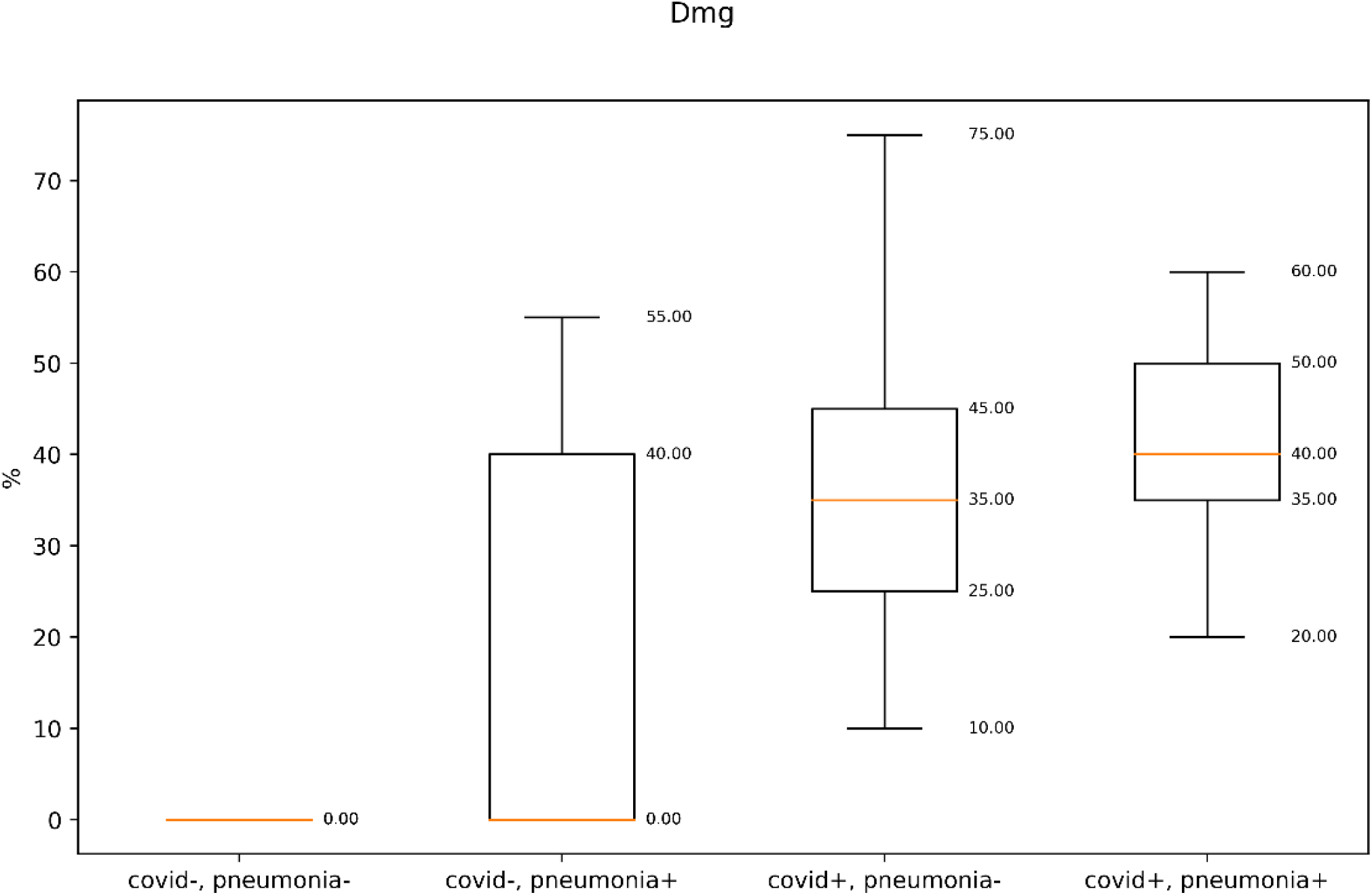
Lung Damage Assessment.

**Appendix A. Figure 2.**
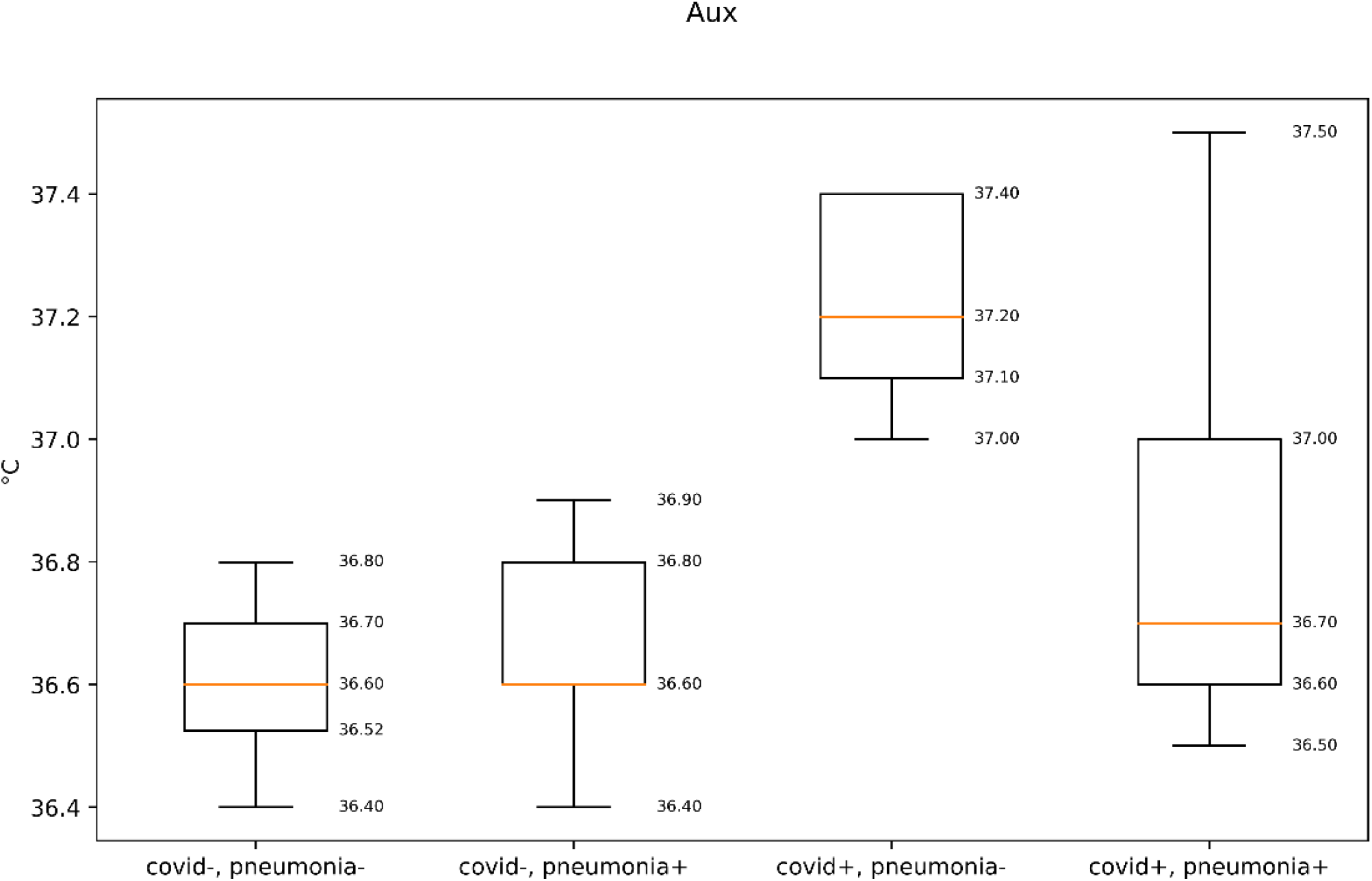
Auxiliary Temperature.

**Appendix A. Figure 3.**
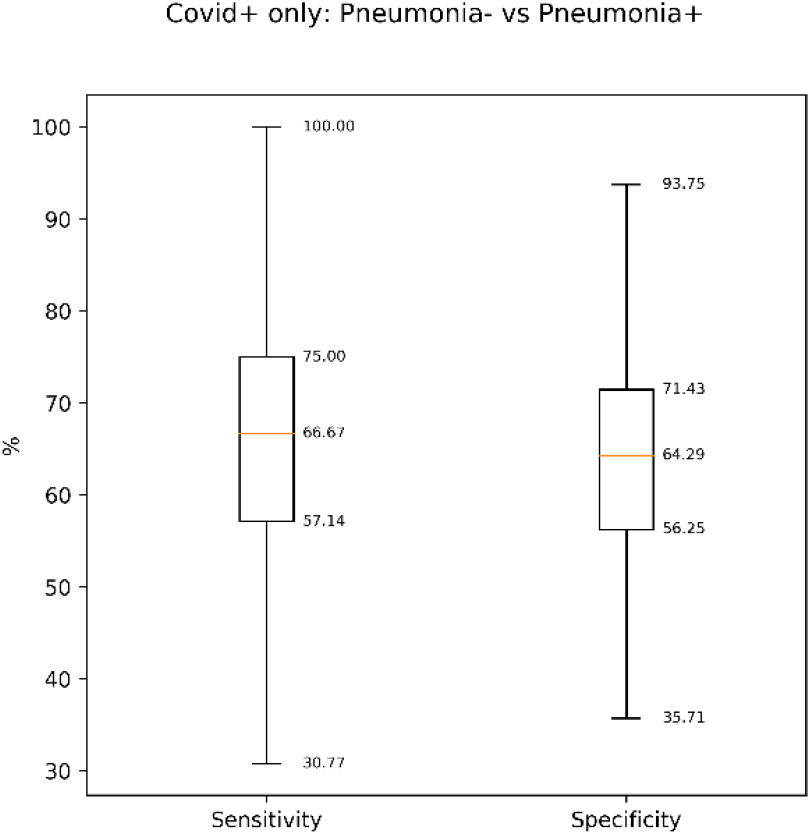
Most valuable metrics: SP (three aggregates), RI max, Int median, Sk median, Distributions:

**Appendix A. Figure 4.**
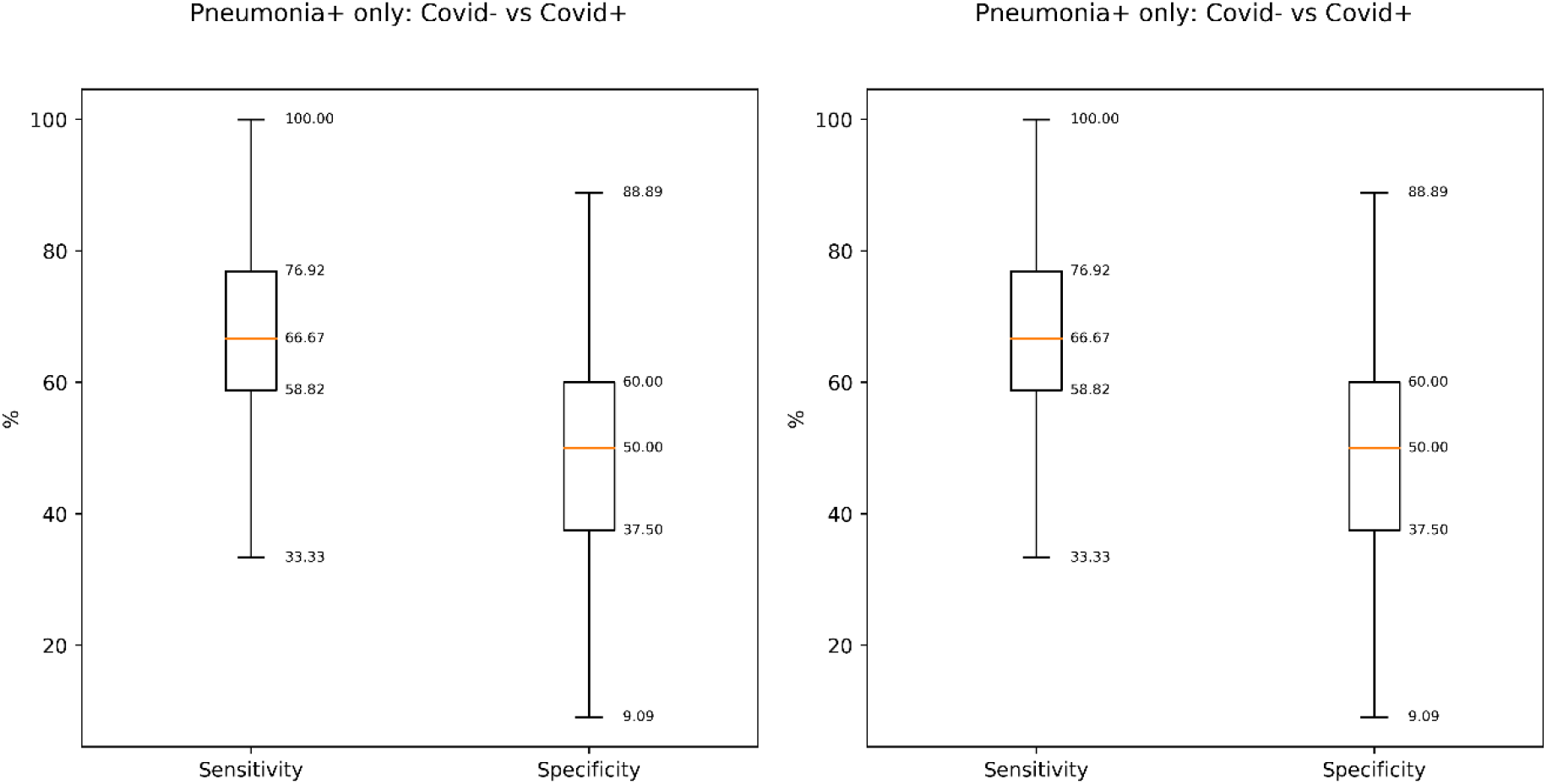
Most valuable metrics: SP (three aggregates), RI max, Int median, Sk median, Distributions:

**Appendix A. Figure 5.**
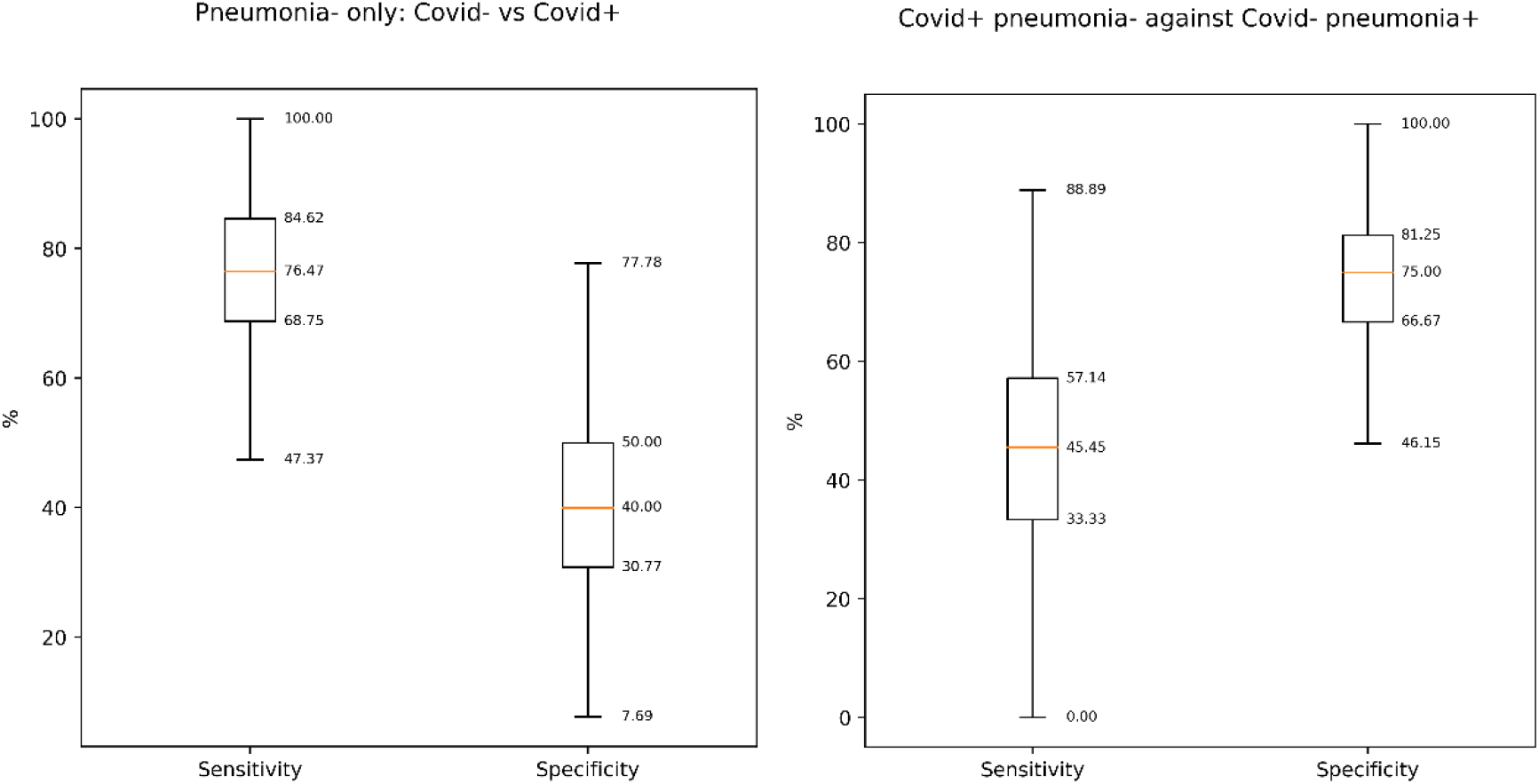
Most valuable metrics: SP (three aggregates), RI max, Int median, Sk median, Distributions:

**Appendix A. Fig.6.**
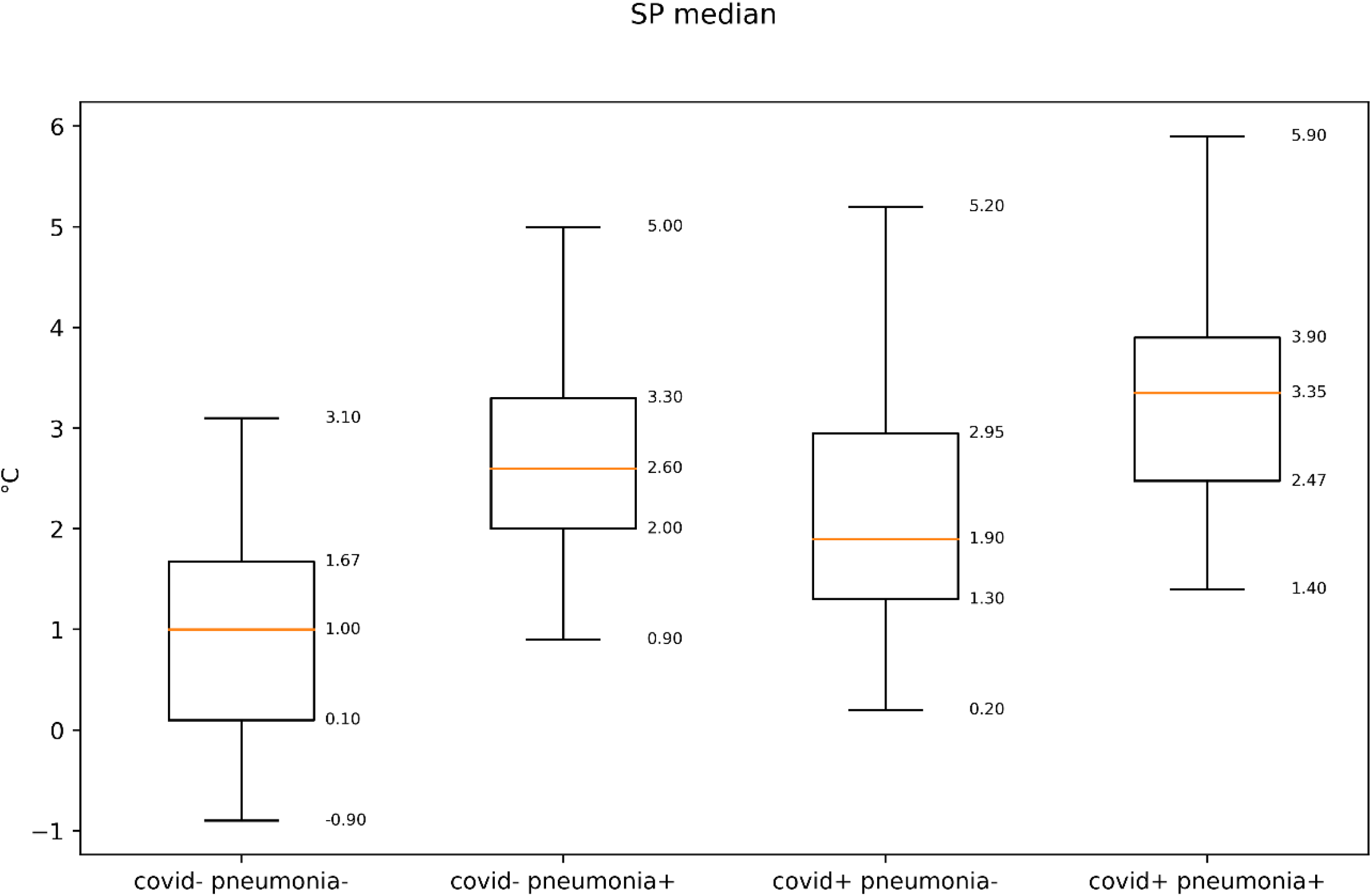
Correlations with side measurements were built to validate if there is higher correlation with specific factor (SpO2, CT, Aux. Temp). SP median. R2 = 0.25 for SpO2 and CT, and 0.08 for Aux. Temp

**Appendix A. Fig.7.**
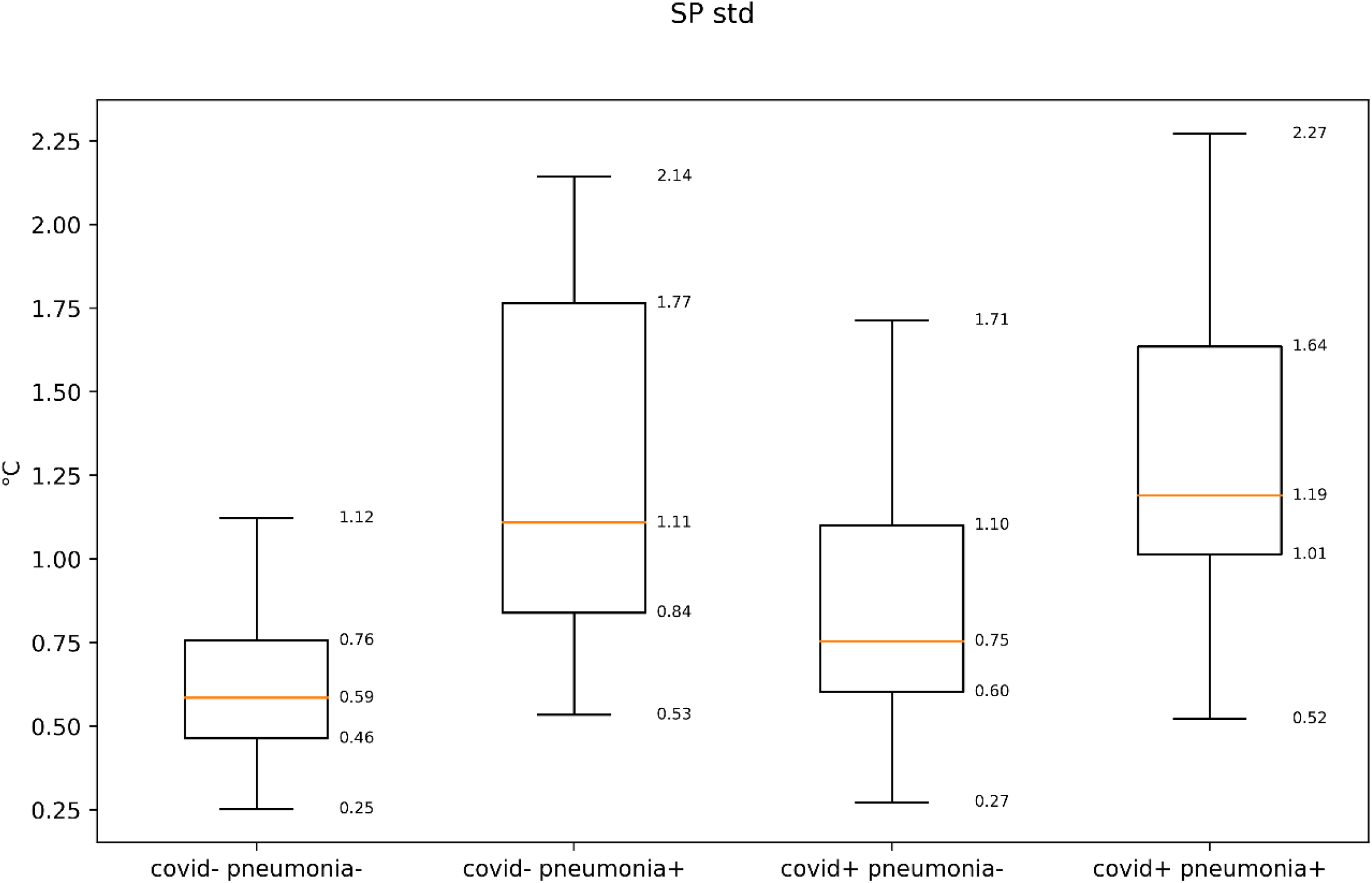
Correlations with side measurements were built to validate if there is higher correlation with specific factor (SpO2, CT, Aux. Temp). SP median. R2 = 0.25 for SpO2 and CT, and 0.08 for Aux. Temp

**Appendix A. Fig.8.**
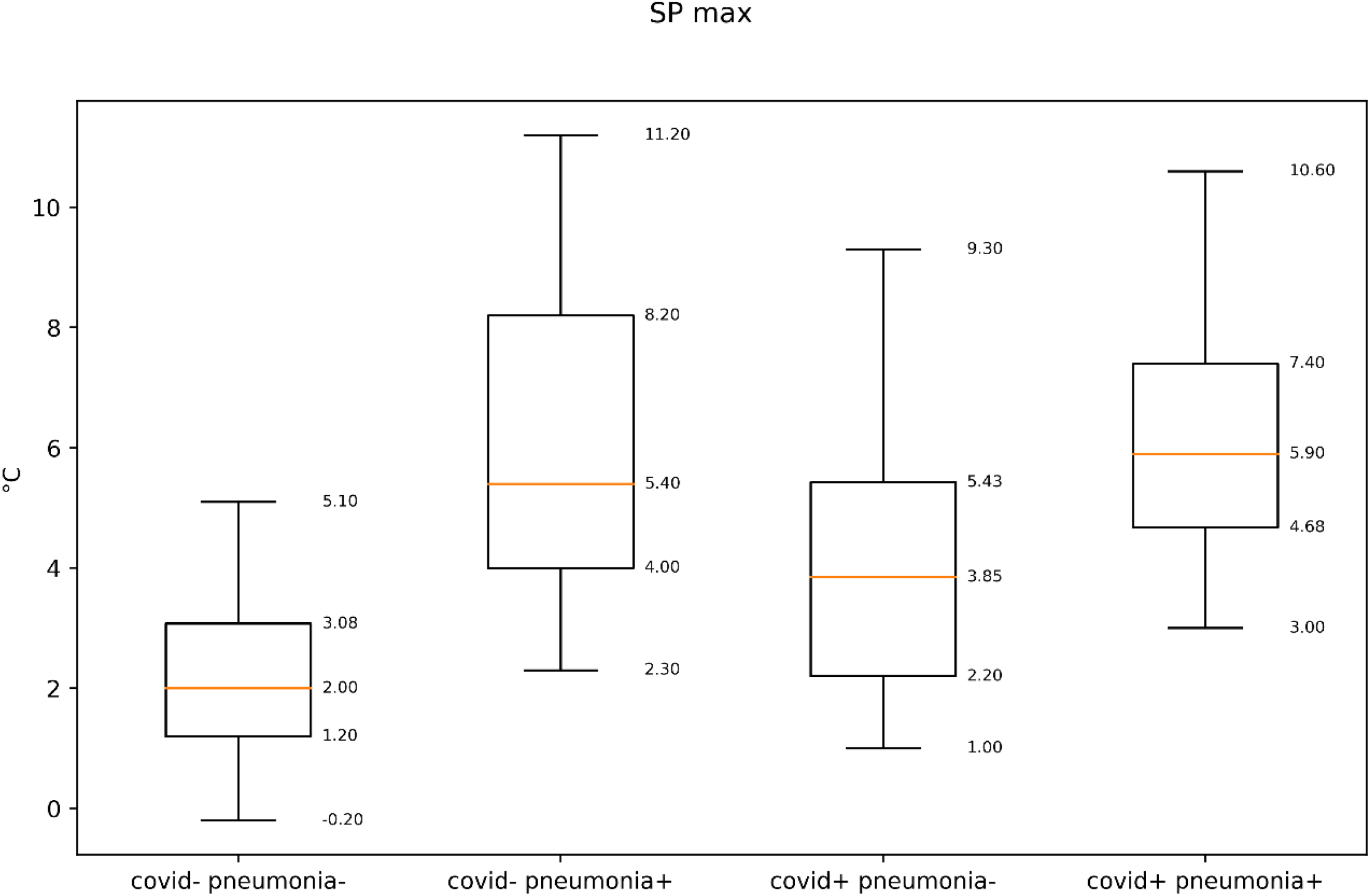
Correlations with side measurements were built to validate if there is higher correlation with specific factor (SpO2, CT, Aux. Temp). SP median. R2 = 0.25 for SpO2 and CT, and 0.08 for Aux. Temp

**Appendix A. Fig.9.**
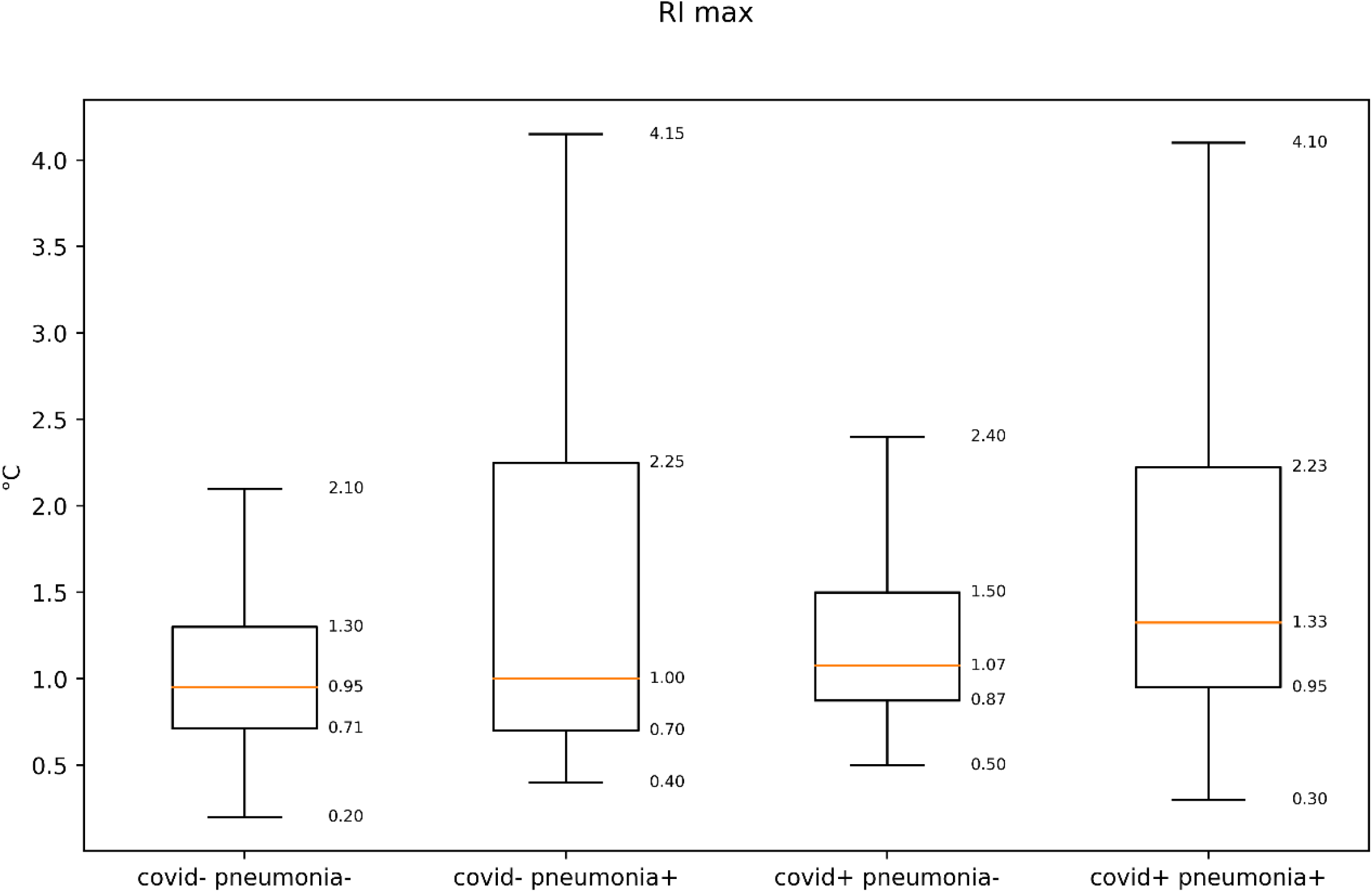
Correlations with side measurements were built to validate if there is higher correlation with specific factor (SpO2, CT, Aux. Temp). SP median. R2 = 0.25 for SpO2 and CT, and 0.08 for Aux. Temp

**Appendix A. Fig.10.**
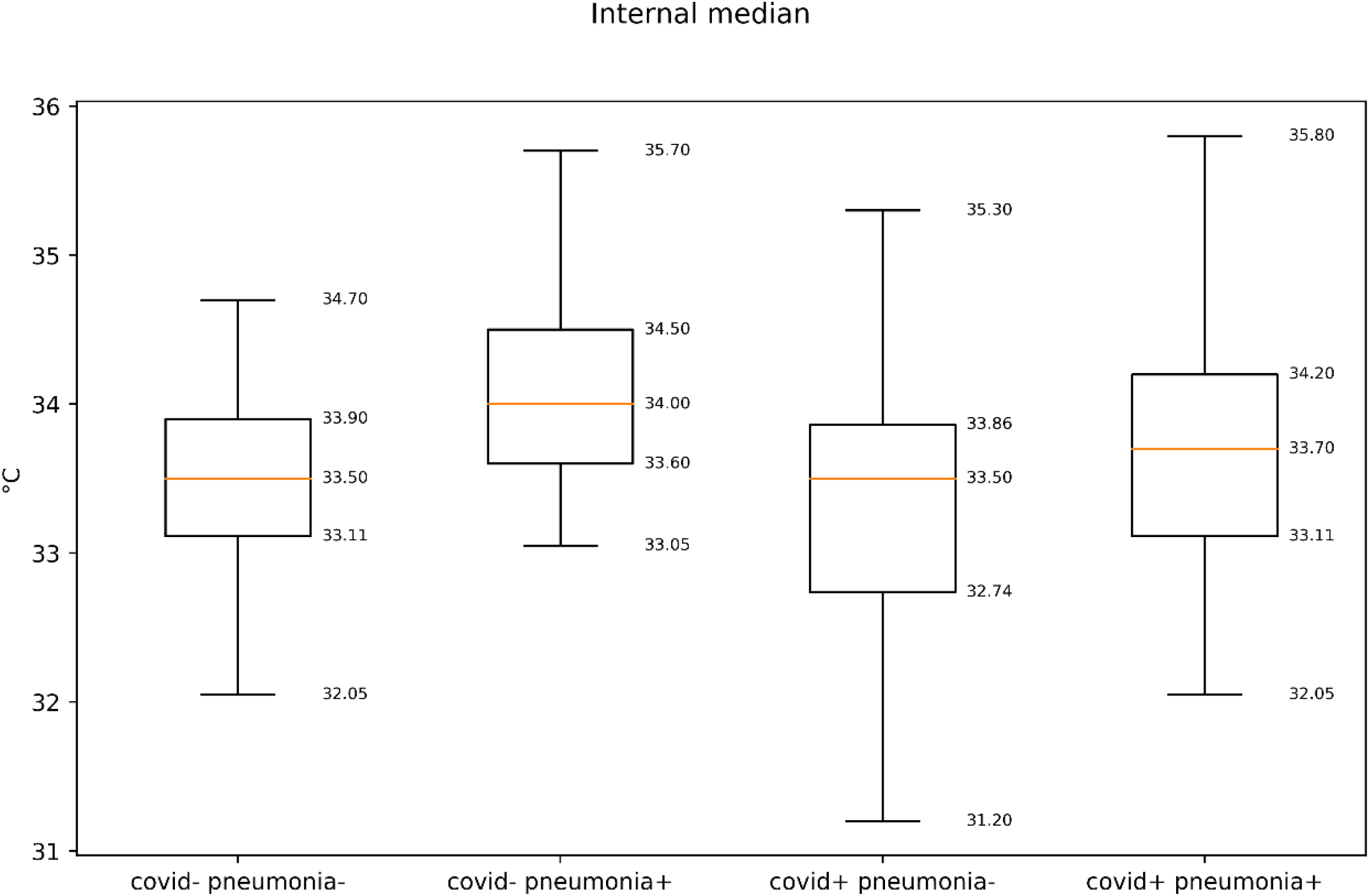
Correlations with side measurements were built to validate if there is higher correlation with specific factor (SpO2, CT, Aux. Temp). SP median. R2 = 0.25 for SpO2 and CT, and 0.08 for Aux. Temp

**Appendix A. Fig.11.**
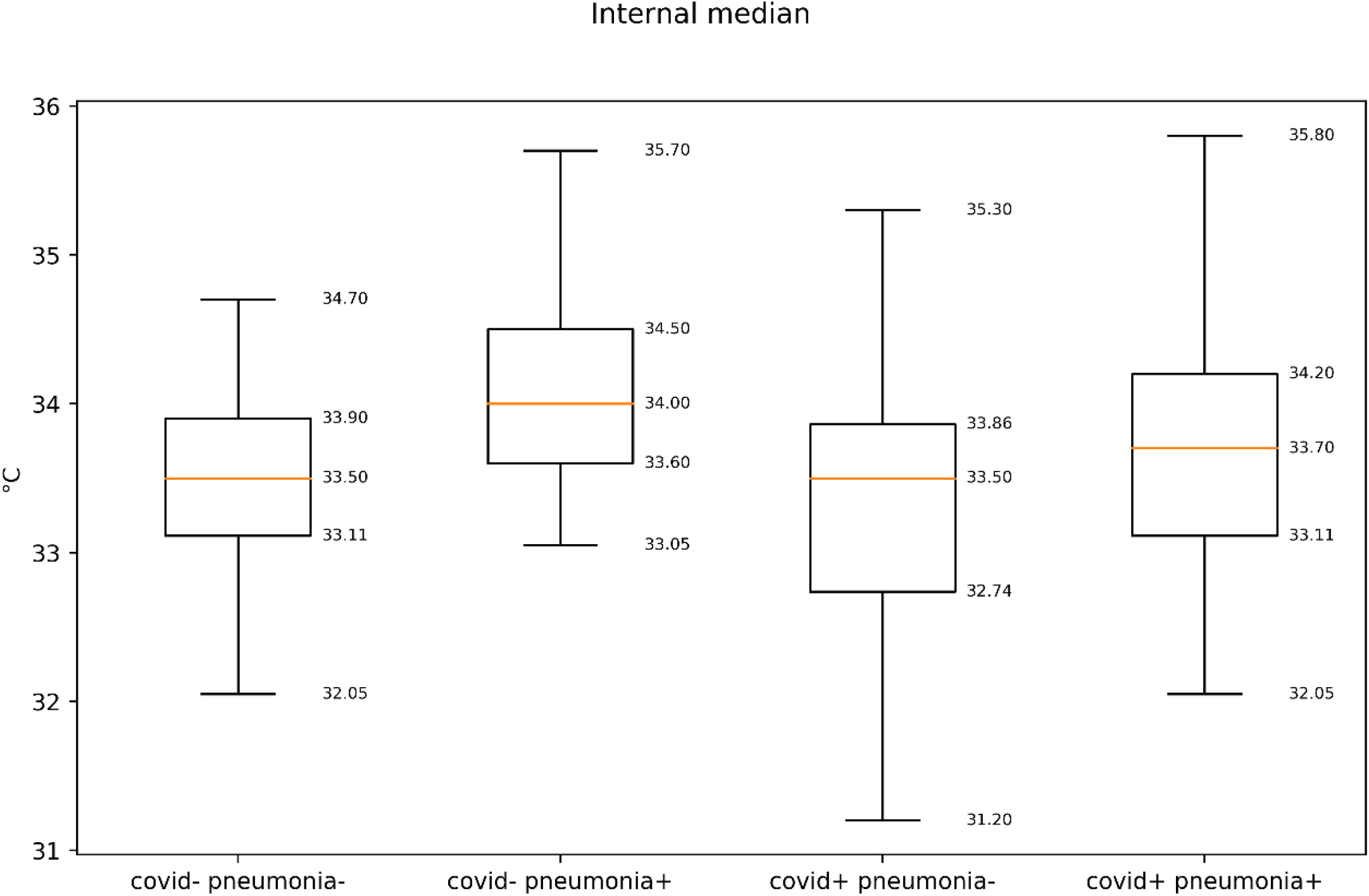
Correlations with side measurements were built to validate if there is higher correlation with specific factor (SpO2, CT, Aux. Temp). SP median. R2 = 0.25 for SpO2 and CT, and 0.08 for Aux. Temp

**Appendix A. Fig.12.**
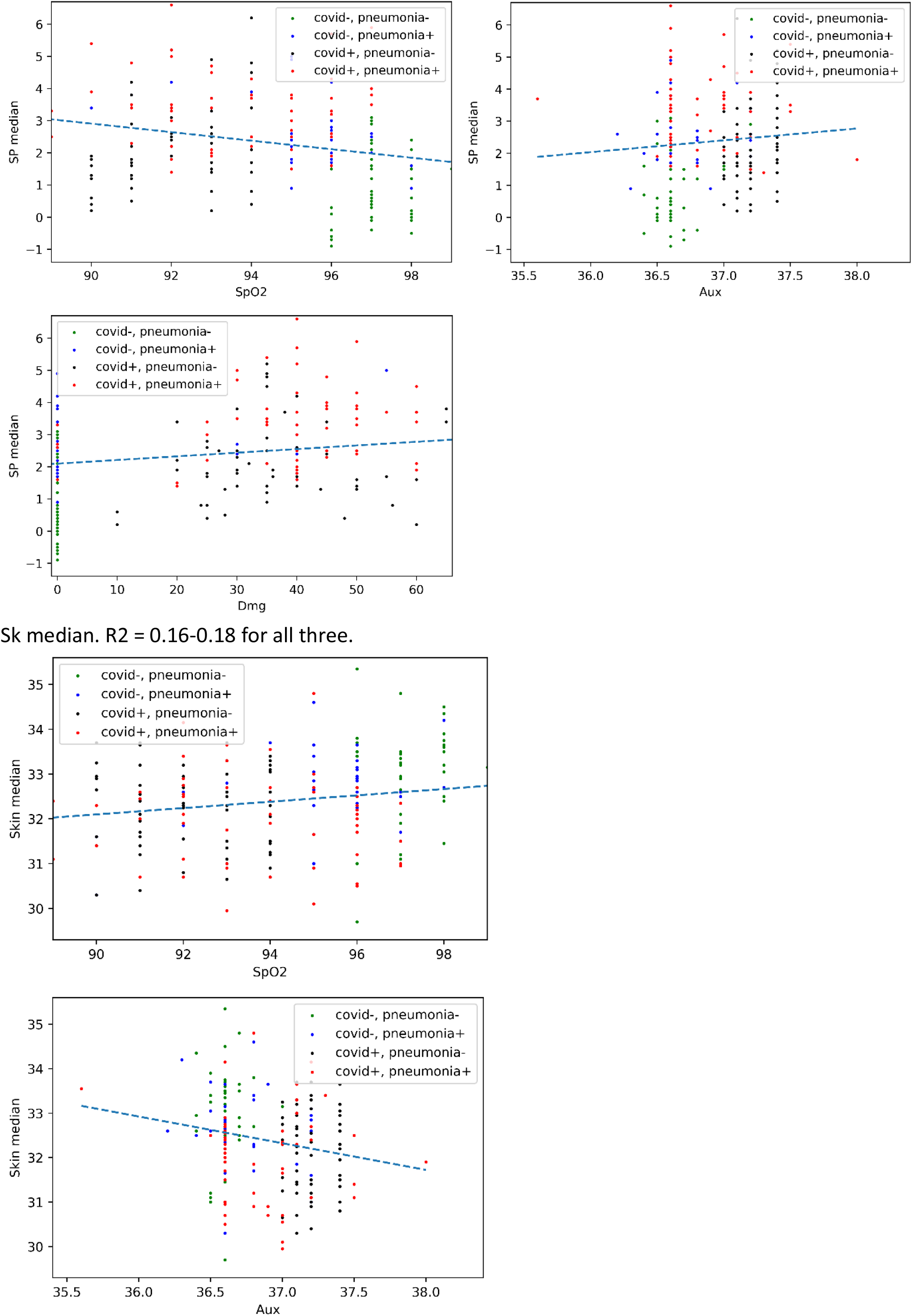
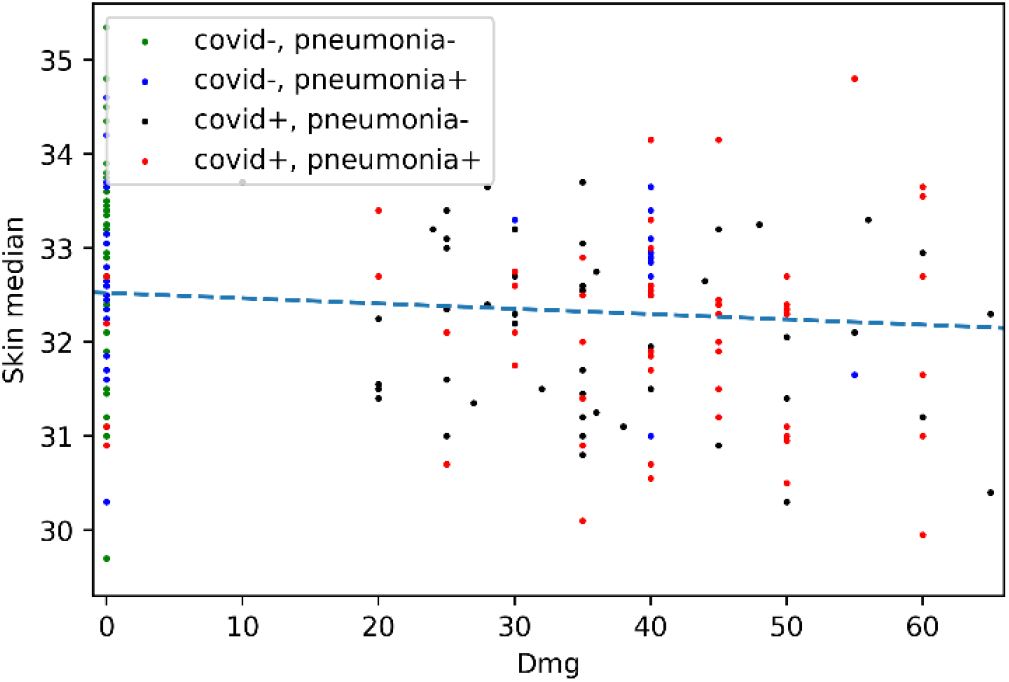
Correlations with side measurements were built to validate if there is higher correlation with specific factor (SpO2, CT, Aux. Temp). SP median. R2 = 0.25 for SpO2 and CT, and 0.08 for Aux. Temp

### Tukey’s multi-comparison method

See https://en.wikipedia.org/wiki/Tukey's_range_test, https://pythonhealthcare.org/2018/04/13/55-statistics-multi-comparison-with-tukeys-test-and-the-holm-bonferroni-method/

This method tests at P<0.05 (correcting for the fact that multiple comparisons are being made which would normally increase the probability of a significant difference being identified). A results of ‘reject = True’ means that a significant difference has been observed.

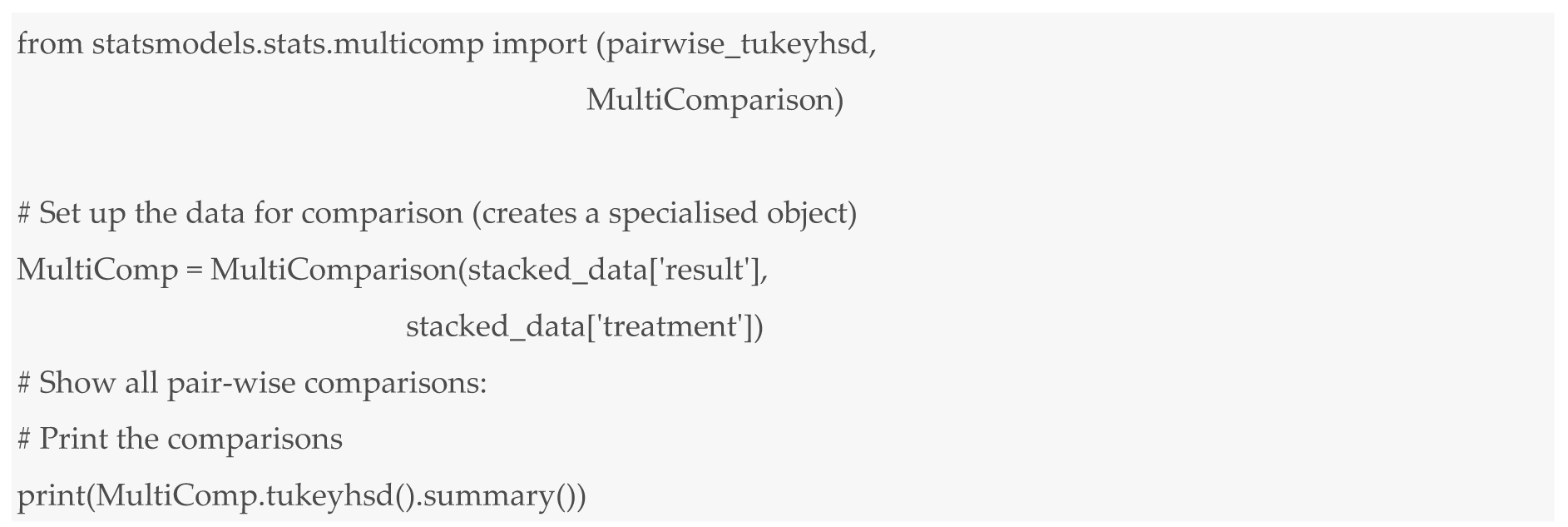

Appendix B. Python code

